# Association analysis between an epigenetic alcohol risk score and blood pressure

**DOI:** 10.1101/2024.02.29.24303545

**Authors:** Helena Bui, Amena Keshawarz, Mengyao Wang, Mikyeong Lee, Scott M. Ratliff, Lisha Lin, Kira S. Birditt, Jessica D. Faul, Annette Peters, Christian Gieger, Thomas Delerue, Sharon L. R. Kardia, Wei Zhao, Xiuqing Guo, Jie Yao, Jerome I. Rotter, Yi Li, Xue Liu, Dan Liu, Juliana F. Tavares, Gökhan Pehlivan, Monique M.B. Breteler, Irma Karabegovic, Carolina Ochoa-Rosales, Trudy Voortman, Mohsen Ghanbari, Joyce B.J. van Meurs, Mohamed Kamal Nasr, Marcus Dörr, Hans J. Grabe, Stephanie J. London, Alexander Teumer, Melanie Waldenberger, David R. Weir, Jennifer A. Smith, Daniel Levy, Jiantao Ma, Chunyu Liu

**Affiliations:** Population Sciences Branch, Division of Intramural Research, National Heart, Lung, and Blood Institute, National Institutes of Health, Bethesda, MD, USA; Framingham Heart Study, Framingham, MA, USA; Department of Biostatistics, Boston University School of Public Health, Boston, MA; Epidemiology Branch, National Institute of Environmental Health Sciences, National Institutes of Health, Department of Health and Human Services, Research Triangle Park, NC, USA; Department of Epidemiology, School of Public Health, University of Michigan, Ann Arbor, MI; Survey Research Center, Institute for Social Research, University of Michigan, Ann Arbor, MI; Institute of Epidemiology, Helmholtz Munich, German Research Center for Environmental Health, German; Institute for Medical Informatics, Biometrics and Epidemiology, Ludwig-Maximilians-Universität München, Munich, Germany; German Center for Cardiovascular Research (DZHK), Partner Site Munich Heart Alliance, Munich, Germany; Research Unit Molecular Epidemiology, Institute of Epidemiology, Helmholtz Munich, German Research Center for Environmental Health, Bavaria, Germany; The Institute for Translational Genomics and Populations, Department of Pediatrics, Lundquist Institute for Biomedical Innovation at Harbor-UCLA Medical Center, Torrance, CA 90502, USA; Population Health Sciences, German Center for Neurodegenerative Diseases (DZNE), Bonn, Germany; Institute for Medical Biometry, Informatics and Epidemiology (IMBIE), Faculty of Medicine, University of Bonn, Bonn, Germany; Department of Epidemiology, Erasmus MC University Medical Center, Rotterdam, The Netherlands; Centro de Vida Saludable de la Universidad de Concepción, Concepción, Chile; Division of Human Nutrition and Health, Wageningen University and Research, Wageningen, The Netherlands; Department of Internal Medicine, Erasmus MC University Medical Center, Rotterdam, The Netherlands; Institute for Community Medicine, University Medicine Greifswald, Greifswald, Germany; German Center for Cardiovascular Research (DZHK), Partner Site Greifswald, Greifswald, Germany; Department of Internal Medicine B, University Medicine Greifswald, Greifswald, Germany; Department of Psychiatry and Psychotherapy, University Medicine, Greifswald, Germany; German Center of Neurodegenerative Diseases (DZNE), Rostock/Greifswald, site Greifswald, Germany; Department of Population Medicine and Lifestyle Diseases Prevention, Medical University of Bialystok, Bialystok, Poland; Gerald J. and Dorothy R. Friedman School of Nutrition Science and Policy, Tufts University, Boston, MA, USA

**Keywords:** epigenetic risk score, DNA methylation, blood pressure, hypertension, alcohol

## Abstract

**Background:** Epigenome-wide association studies have revealed multiple DNA methylation sites (CpGs) associated with alcohol consumption, an important lifestyle risk factor for cardiovascular diseases.

**Results:** We generated an alcohol consumption epigenetic risk score (ERS) based on previously reported 144 alcohol-associated CpGs and examined the association of the ERS with systolic blood pressure (SBP), diastolic blood pressure (DBP), and hypertension (HTN) in 3,898 Framingham Heart Study (FHS) participants. We found an association of alcohol intake with the ERS in the meta-analysis with 0.09 units higher ERS per drink consumed per day (*p* < 0.0001). Cross-sectional analyses in FHS revealed that a one-unit increment of the ERS was associated with 1.93 mm Hg higher SBP (*p* = 4.64E-07), 0.68 mm Hg higher DBP (*p* = 0.006), and an odds ratio of 1.78 for HTN (*p* < 2E-16). Meta-analysis of the cross-sectional association of the ERS with BP traits in eight independent external cohorts (n = 11,544) showed similar relationships with blood pressure levels, i.e., a one-unit increase in ERS was associated with 0.74 (*p* = 0.002) and 0.50 (*p* = 0.0006) mm Hg higher SBP and DBP, but could not confirm the association with hypertension. Longitudinal analyses in FHS (n = 3,260) and five independent external cohorts (n = 4,021) showed that the baseline ERS was not associated with a change in blood pressure over time or with incident HTN.

**Conclusions:** Our findings provide proof-of-concept that utilizing an ERS is a useful approach to capture the recent health consequences of lifestyle behaviors such as alcohol consumption.

## Introduction

Approximately 178,307 people die annually from alcohol-related causes, making alcohol consumption one of leading preventable causes of death in the United States[1]. Alcohol has complex effects on multiple biological processes and systems, including the cardiovascular system. Several studies suggest that habitual, heavy alcohol use can lead to cardiovascular sequelae such as dilated cardiomyopathy and heart failure[2]. However, the benefits and potential arms of moderate drinking have been a subject of controversy. A few studies have indicated that the association presented a J-shaped curve between alcohol consumption and a lower risk of cardiovascular disease[3–6]. While studies with Mendelian randomization method suggested a non-linear and increased risk of cardiovascular risks with any dose of alcohol intake[3,5]. Additionally, multiple studies support a causal relationship of alcohol consumption with elevated blood pressure and the risk of hypertension (HTN)[2,7]. Furthermore, high blood pressure is one of the leading risk factors for cardiovascular diseases (CVDs)[8,9]. Therefore, understanding the molecular changes underlying alcohol consumption is crucial to comprehend the relationship between alcohol consumption, high blood pressure, and CVD.

One of the most studied epigenetic modifications, DNA methylation, regulates gene expression through the transfer of a methyl group onto DNA cytosine-phosphate-guanine (CpG) sites. The extent of DNA methylation at certain CpG sites is associated with phenotypic variation in numerous CVD-related traits including body mass index (BMI)[10], blood lipids[11], glycemic traits[12], blood pressure[13], and inflammatory biomarkers[14]. DNA methylation has also been linked to lifestyle behaviors such as alcohol consumption. A large-scale meta-analysis of data from thirteen population-based cohorts including the Framingham Heart Study (FHS) identified 144 differentially methylated CpG sites associated with heavy alcohol intake[15].

A standardized, biomarker of alcohol consumption may correct for limitations of self-reported alcohol consumption, such as impression management bias[16] or faulty recall of drinking history[17,18], and reveal alcohol-related disease risks that otherwise might not be apparent. In this study, we used 144 alcohol-related, differentially-methylated CpGs[15] to generate an alcohol consumption epigenetic risk score (ERS) and examine the association of the ERS with blood pressure traits in cross-sectional and longitudinal analyses. We hypothesized that a DNA methylation-based alcohol consumption ERS would be associated with blood pressure, cross-sectionally and longitudinally. We tested our hypothesis by analyzing the association of our alcohol-associated ERS with blood pressure traits, including systolic blood pressure (SBP), diastolic blood pressure (DBP), and HTN in 3,898 FHS participants. In addition, we carried out replication analyses of these findings in eight independent cohorts using meta-analysis (**Figure 1**). The alcohol consumption ERS provides an opportunity to investigate the relations of alcohol intake to health outcomes in situations where self-reported intake data is unavailable or unreliable.

**Figure 1.**
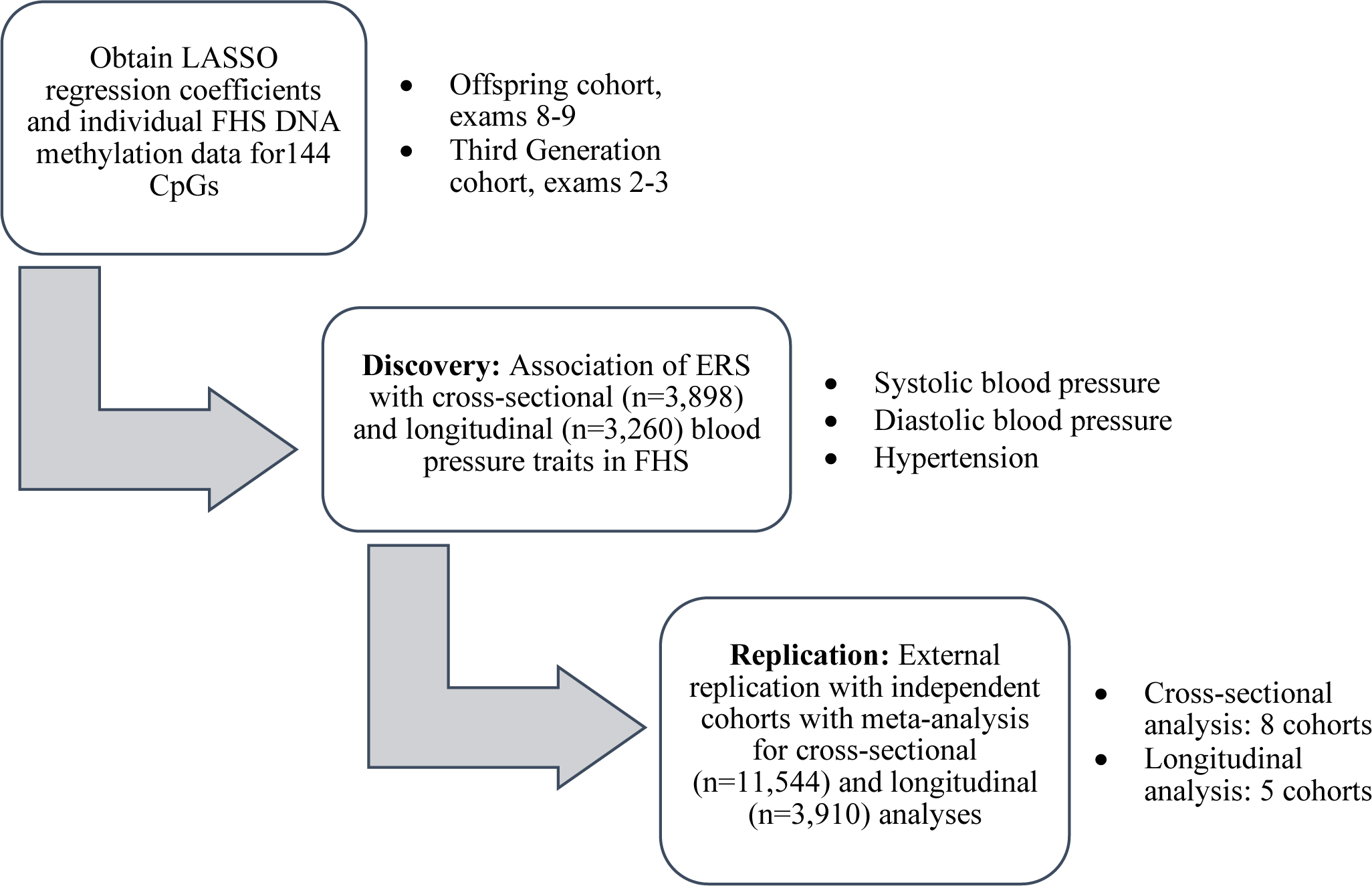
Study design.

## METHODS

### Study population

Data from nine population-based cohort studies were used in the analysis. In addition to the FHS[19], our investigation included the Agricultural Lung Health Study (ALHS)[20], the Cooperative Health Research in the Region Augsburg (KORA)[21], the Genetic Epidemiology Network on Arteriopathy (GENOA) Study[22], the Health and Retirement Study (HRS)[23], the Multi-Ethnic Study of Atherosclerosis (MESA) Study[24], the Rhineland Study[25], the Rotterdam Study[26], and the Study of Health in Pomerania (SHIP)[27]. Institutional review committees of all cohorts approved this study, and all study participants provided written informed consent. In each cohort, participants with prevalent CVD (angina pectoris, coronary insufficiency, cerebrovascular accident, atherothrombotic infarction of the brain, transient ischemic attack, cerebral embolism, intracerebral hemorrhage, subarachnoid hemorrhage, or intermittent claudication), prevalent heart failure, and prevalent atrial fibrillation were excluded. We also excluded participants without DNA methylation data at baseline examination at which blood samples were collected for DNA methylation profiling. After exclusions, 3,898 participants in FHS and 11,544 participants in eight independent external cohorts were included in cross-sectional association analyses, while 3,260 participants in FHS and 3,910 participants in five external cohorts were included in longitudinal association analyses (**Figure 1**).

### Clinical and behavioral data collection

Overall, clinical data for traits such as age, BMI, SBP, DBP, and the use of antihypertensive medication were collected at in-person examinations. HTN was defined as SBP ≥ 140 mm Hg, DBP ≥ 90 mm Hg, or use of antihypertensive medication for treating hypertension at the examination. We added 15 mm Hg and 10 mm Hg to a participant’s measured SBP and DBP values, respectively, for participants currently using antihypertensive medication.

Participants’ cigarette smoking status was determined based on self-reported smoking behavior. Current smokers were defined as participants who smoked on average at least one cigarette per day in the past year; former smokers were defined as participants who previously smoked on average at least one cigarette per day but stopped smoking for at least one year; never smokers were defined as participants who never smoked. Self-reported alcohol intake was captured via questionnaires wherein the participants reported the frequency with which they consumed various alcoholic beverages (i.e., beer, liquor, or wine). This study included nine population-based cohorts, and therefore, we focused on habitual alcohol consumption in general populations rather than examining specifically for alcohol disorder. Study-specific methods for clinical data collection are presented in the **Supplemental Text** and **Supplemental Table 1**.

### DNA methylation data collection and processing

Whole blood samples were assayed for DNA methylation via the Infinium Human Methylation 450 BeadChip platform or Infinium MethylationEPIC platform (San Diego, CA) (**Supplemental Text**). The methylated probe intensity and total probe intensities were extracted using the Illumina Genome Studio (version 2011.1) with the methylation module (version 1.9.0). Preprocessing of the methylated (M) signal and unmethylated signal (U) was conducted; methylation beta-value (β_M_) was defined as 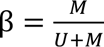. Further information regarding DNA extraction and processing has been outlined[15]. Further information regarding the collection of DNA samples and assay details about each external cohort are presented in the Supplemental Materials.

### Epigenetic risk score calculation

To develop the ERS for this investigation, we used 144 CpGs identified in a previous EWAS meta-analysis[15]. The score was calculated by multiplying individual-level methylation values at each CpG site by the CpG’s corresponding Least Absolute Shrinkage and Selection Operator (LASSO) β estimate. LASSO is a regression analysis method that performs both variable selection and regularization to enhance the prediction accuracy and interpretability of the resulting statistical model by shrinking the coefficients of some variables to exactly zero[28]. The weighted individual values were summed across all 144 CpGs sites to create an ERS for each participant, representing weighted DNA methylation levels in response to alcohol consumption. This methodology was applied to all participating cohorts’ methylation data to generate the ERS. Across five cohorts, one drink of alcohol consumption was associated with 0.09 higher unit of ERS (**Supplemental Table 2**). Methods for calculating the ERS for cohorts missing certain CpG methylation values can be found in the **Supplemental Text**.

### Analysis of the epigenetic risk score with blood pressure traits in FHS (discovery)

We performed both cross-sectional and longitudinal regression analyses in FHS to examine the association between the ERS (independent variable) and blood pressure traits: SBP (continuous), DBP (continuous), and HTN (dichotomous) (dependent variables). Linear mixed regression models were used to evaluate the association of the ERS with the two continuous blood pressure traits. Generalized estimating equations (GEE) were used to evaluate the association of the ERS with dichotomous HTN. A total of 3,898 participants were included in the cross-sectional analysis from the FHS Offspring cohort (n = 2,393; examination 8) and FHS Third Generation cohort (n = 1,505; examination 2) participants. All models were adjusted for age, age-squared, sex, BMI, familial correlation (for family data), and current smoking status.

Longitudinal analyses of all blood pressure traits included FHS Offspring cohort participants (n = 1,932) who attended both examinations 8 and 9 and Third Generation participants (n = 1,328) who attended both examinations 2 and 3. Our linear mixed regression models evaluated the association of change in blood pressure over time (i.e.**, Δ**SBP and **Δ**DBP) with the baseline ERS after adjusting for baseline age, baseline age-squared, sex, baseline BMI, baseline smoking status, baseline SBP/DBP (i.e., if the model’s outcome was **Δ**DBP, we adjusted for baseline DBP), and time between baseline and the follow-up examination. Our GEE models evaluated the association of incident HTN with the baseline ERS after adjusting for baseline age, baseline age-squared, baseline BMI, baseline smoking status, and time between baseline and follow-up examination; in addition, these GEE models excluded all participants with HTN at baseline examination. In the sensitivity analysis, we defined participants with stage 1 hypertension using the 2017 guideline (i.e., ≥ 130/80 SBP/DBP mm Hg or with antihypertension treatment)[29]. We performed cross-sectional and longitudinal GEE models to investigate the associations of ERS with prevalent and incident hypertension using the new definition.

### Meta-analysis (replication)

For replication, independent external cohorts (n = 11,544) were used in an inverse-variance weighted, fixed-effects meta-analysis by assuming that there is one true effect between ERS and a BP trait. Eight studies were used in cross-sectional meta-analysis for replication while five studies were utilized in the longitudinal meta-analysis (n = 4,021).

### Analysis of the epigenetic risk score with blood pressure traits in participants without antihypertension medication

To minimize the possible effects of antihypertension medication on DNA methylation, we conducted a sensitivity-analysis among participants without antihypertension medication in five cohorts (i.e., FHS, GENOA, HRS, Rhineland Study, and SHIP). Similar to the primary analysis, we conducted the cross-sectional analysis using linear mixed effects model with ERS as the independent variable and blood pressure traits as the dependent variables in each cohort.

### Analysis of the epigenetic risk score with alcohol consumption

We used a linear mixed regression model to test the cross-sectional association between the ERS (outcome) and self-reported alcohol intake (exposure) in each of the five cohorts (i.e., FHS, GENOA, HRS, Rhineland Study, and SHIP). The change in the ERS associated with one drink of alcohol consumption per day was calculated with adjustment for age, age-squared, sex, BMI, current smoking status, and familial correlation.

### Association analysis of alcohol consumption with blood pressure traits

To compare the association of blood pressure traits with ERS and questionnaire-based alcohol consumption, we performed cross-sectional (i.e., FHS, GENOA, HRS, Rhineland Study, and SHIP) and longitudinal (i.e., FHS, GENOA, and SHIP) analyses between blood pressure traits and alcohol consumption. We used linear mixed or GEE models to quantify the associations between SBP/DBP/HTN (outcome variables) and alcohol consumption (predictor). Covariates included age, age-squared, sex, BMI, current smoking status, and family structure.

### Association of epigenetic risk score with biological biomarkers of alcohol intake

We tested the association of the ERS with two established biomarkers of chronic alcohol consumption: aspartate amino transferase (AST) and alanine aminotransferase (ALT) concentrations. Separate linear mixed regression models were used with each enzyme as the dependent variable. Serum AST and ALT were measured on fasting morning samples using the kinetic method (Beckman Liquid-State Reagent Kit)[30]. Model 1 (i.e., the reduced model) quantified the association between the self-reported alcohol intake and liver enzyme concentrations after adjusting for age, sex, BMI, and smoking status. Model 2 (i.e., the full model) further adjusted for the ERS. In order to compare the two models, we also performed a likelihood ratio test (LRT) to gauge whether the addition of the ERS significantly improved model fit.

### Analysis of individual alcohol-associated CpGs with blood pressure traits in FHS

We examined the cross-sectional association of 144 DNA methylation probes from ERS with blood pressure traits in the FHS. We applied the linear mixed effect model to account for the pedigree with each CpG probe as the predictor variable and SBP/DBP as the outcome variable. Covariates included age, age-squared, sex, BMI, and current smoking status.

All statistical analyses were conducted using the R (version 4.0.3) software package[31]. Meta-analyses was conducted with the *metafor* package (version 3.0.2)[32]. LRT was performed using the ‘lrtest’ function in the R package *lmtest* in R (version 0.9.39)[33]. Statistical significance was defined as two-sided p<0.05.

## RESULTS

### Participant characteristics

This present study included discovery and replication analyses (**Figure 1**). The discovery association analysis was performed in the FHS. The replication association analysis and meta-analysis was performed in up to eight cohorts (**Figure 1**). At the baseline examination, FHS participants (n = 3,898) were, on average, 58 years old (SD = 13 years) and consisted of slightly more women (55%) than men (45%) (**Supplemental Table 3**). In addition, approximately 42% of FHS participants had hypertension at baseline. Furthermore, women consumed less alcohol compared to men (mean alcohol intake 0.3 drinks/day vs. 0.7 drinks/day). The FHS participants (n = 3,260) were followed up for six years and used for longitudinal association analyses with blood pressure traits (**Supplemental Table 2**).

Overall, mean age of participants in the eight independent cohorts (ntotal = 11,544) ranged from of 49 years (SHIP) to 68 years (HRS) (**Supplemental Tables 3-5**). Similar to what was observed in FHS, women reported a lower average amount of alcohol consumed per day compared to men. DNA methylation was measured using blood samples collected at the same time when alcohol consumption data were assessed in all nine cohorts. Blood pressure traits measured contemporaneously were used in cross-sectional analysis and those measured six to ten years after alcohol consumption measurement were used for longitudinal studies (**Supplemental Table 1**). Additionally, the mean values of the ERS ranged from −15.35 (SD = 0.74) to −3.85 (SD = 0.61) across all cohorts at the baseline examination (**Supplemental Table 6**).

### Epigenetic risk score and blood pressure: cross-sectional and longitudinal analysis in FHS

The alcohol intake showed a significant association with ALT (*p* = 2.9E-09) and AST (*p* = 1.3E-10), but borderline for the AST/ALT ratio (*p* = 0.054) in Model 1. The addition of the ERS in Model 2 (i.e., full model) improved model fit with respect to cross-sectional ALT (*p* = 2.8E-07), AST (*p* = 2.3E-12), and the AST/ALT ratio (*p* = 0.0076) (**Supplemental Table 7**).

Cross-sectional analyses in FHS participants revealed significant association of the ERS with SBP, DBP, and HTN. A one-unit increment of the ERS was associated with a 1.98 mm Hg higher SBP (SE = 0.39, *p* = 4.6E-07), a 0.68 mm Hg higher DBP (SE = 0.25, *p* = 0.006), and an odds ratio of 1.78 for HTN (95% CI = [1.55, 2.04], *p* < 2E-16) (**Table 1**). In contrast, longitudinal analyses did not reveal significant associations of the ERS with ΔSBP, ΔDBP, or incident HTN (*p* > 0.3 for all) (**Table 1**).

**Table 1.** Cross-sectional and longitudinal association analyses of epigenetic risk score and blood pressure traits in the Framingham Heart Study.

| **Continuous variable** | **$\beta$** | **SE** | **p** |
| --- | --- | --- | --- |
| SBP | 1.98 | 0.39 | 4.64E-07 |
| DBP | 0.68 | 0.25 | 0.006 |
| ΔSBP | 0.44 | 0.44 | 0.32 |
| ΔDBP | -0.0012 | 0.26 | 0.99 |
| **Binary variable** | **OR** | **95% CI** | **p** |
| Hypertension | 1.78 | 1.55, 2.04 | < 2E-16 |
| Incident hypertension | 1.02 | 0.83, 1.24 | 0.85 |

SBP/DBP, systolic/diastolic blood pressure; ΔSBP/ ΔDBP, longitudinal change in systolic/diastolic blood pressure; SE, standard error; OR, Odds ratio; 95% CI, 95% confidence interval. The cross-sectional and longitudinal analyses were performed on 3,898 and 3,260, respectively, Framingham Heart Study (FHS) participants.

In the sensitivity analysis with the updated definition for stage 1 hypertension (i.e., ≥ 130/80 mm Hg or with the antihypertension treatment), we observed consistent results as the stage 2 hypertension definition (i.e., ≥ 140/90 mm Hg or with the antihypertension treatment). One-unit higher ERS was positively associated with the prevalent hypertension (OR = 1.70, 95% CI = [1.54, 1.98], *p* < 2E-16) but not significantly associated with the incident hypertension (*p* = 0.98) (**Supplementary Table 8**).

### Replication meta-analysis

Meta-analysis of eight independent external cohorts (n = 11,544) revealed significant cross-sectional associations. A one-unit greater ERS was associated with a 0.74 (95% CI = [0.26, 1.22], *p* = 0.002) mm Hg higher SBP (**Figure 2**) and a 0.50 (95% CI = [0.21, 0.78], *p* = 0.0006) mm Hg higher DBP (**Figure 3**). As can be seen in the forest plots, there was heterogeneity in the meta-analysis results for SBP (Q = 17.27, *p* = 0.008), but not for DBP (Q = 5.16, *p* = 0.52). No significant association was observed between the ERS and HTN in meta-analysis (OR = 1.02, 95% CI = [0.83, 1.24], *p* = 0.10; **Supplemental Figure 1**). As a sensitivity analysis for cross-sectional HTN, meta-analysis was replicated excluding the Rhineland Study, which accounted for 73% of the pooled effect size, but did not change results significantly (**Supplemental Figure 2**).

**Figure 2.**
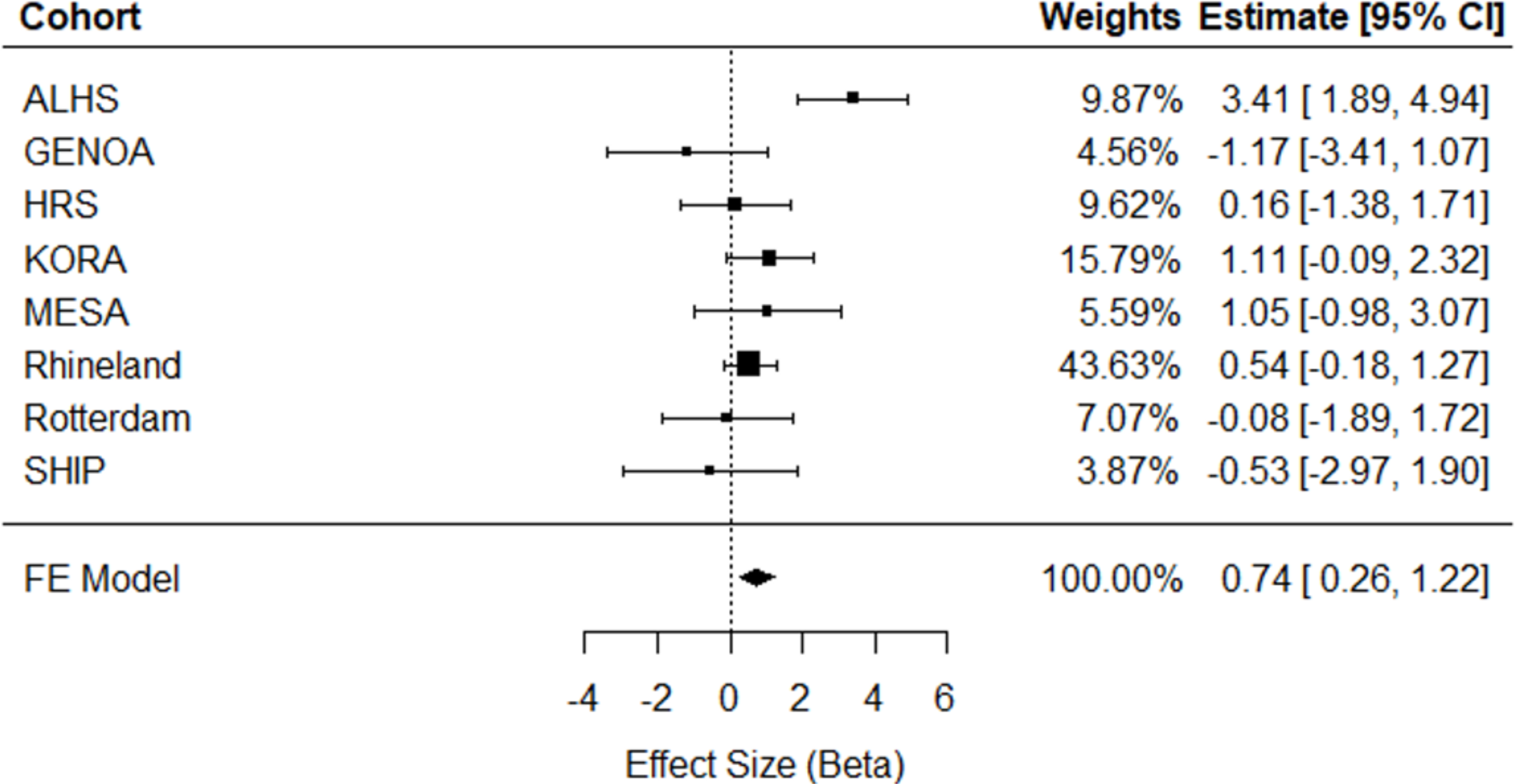
Meta-analysis of cross-sectional association analyses of ERS in relation to systolic blood pressure in eight independent external cohorts (n = 11,544). ALHS, Agricultural Lung Health Study; GENOA, Genetic Epidemiology Network of Arteriopathy; HRS, Health and Retirement Study; KORA, Cooperative Health Research in the Region Augsburg; MESA, Multi-Ethnic Study of Atherosclerosis; SHIP, Study of Health in Pomerania; FE, Fixed Effect; 95% CI, Confidence Interval.

**Figure 3.**
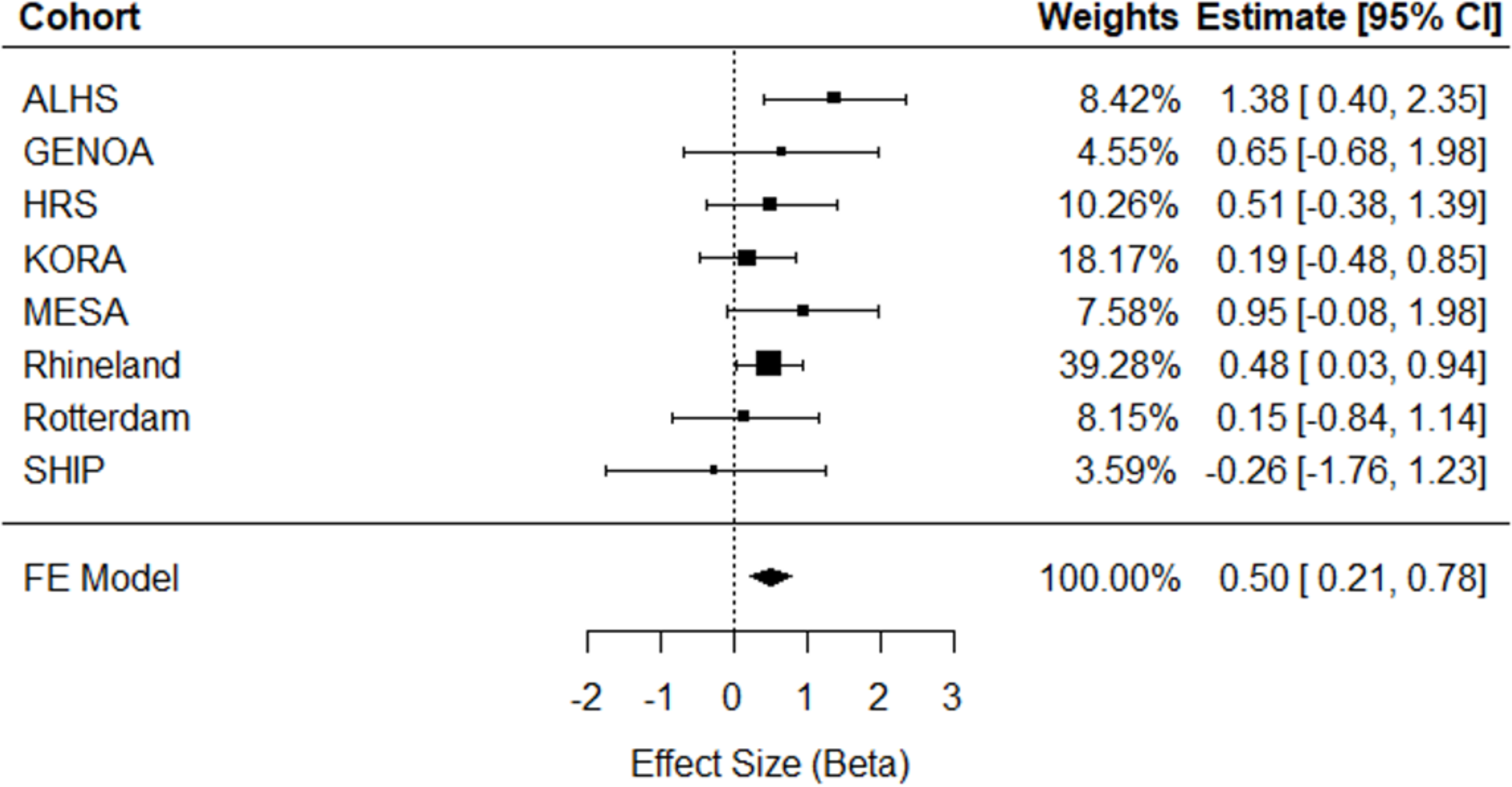
Cross-sectional meta-analysis of ERS in relation to diastolic blood pressure in eight independent external cohorts (n = 11,544). ALHS, Agricultural Lung Health Study; GENOA, Genetic Epidemiology Network of Arteriopathy; HRS, Health and Retirement Study; KORA, Cooperative Health Research in the Region Augsburg; MESA, Multi-Ethnic Study of Atherosclerosis; SHIP, Study of Health in Pomerania; FE, Fixed Effect; 95% CI, Confidence Interval.

No significant associations of the ERS with blood pressure traits in longitudinal meta-analysis were observed (ΔSBP β = 0.63, 95% CI = [-0.12, 1.38], *p* = 0.10; ΔDBP β = 0.10. 95% CI = [-0.30, 0.50], *p* = 0.61; incident HTN β (log OR) = 0.003, 95% CI [-0.05, 0.06], *p* = 0.92). These analyses included five cohorts (GENOA, KORA, MESA, the Rotterdam Study, and SHIP) that had follow-up examination data available (for ΔSBP, ΔDBP total sample size n = 3,910; for incident HTN, n = 3,228). **Supplemental Figures 3, 4, and 5** display the meta-analysis results for ΔSBP, ΔDBP, and incident HTN, respectively. Cross-sectional and longitudinal analysis results for individual cohort are shown in **Supplemental Tables 9and 10**, respectively.

### Sensitivity analyses

#### Association of epigenetic risk score with blood pressure traits in participants without antihypertension medication

We conducted sensitivity analyses between ERS and SBP/DBP in 2577 FHS participants (66.1% of the entire study sample) who were not receiving antihypertension medication. We observed stronger results in the participants without antihypertension treatment compared to the results in all participants (**Supplementary Table 11**). One-unit higher ERS was associated with a 2.6 mm Hg higher SBP (*p* = 1.5E-7) and a 1.54 mm Hg higher DBP (*p* = 1.1E-6), which were stronger than the estimates obtained from all FHS participants (SBP: β =1.98, *p* = 4.6E-7; DBP: β = 0.68, *p* = 0.006). However, associations were not significant between ERS and SBP/DBP among untreated participants (*p* > 0.05) in the four external independent cohorts (**Supplementary Table 11**), except a marginally significant association between ERS and DBP (β = 0.40, *p* = 0.09) in Rhineland untreated participants. Of note, most of the cross-sectional associations were nonsignificant in these four cohorts between ERS and blood pressure traits before treated participants were removed (**Supplementary Table 9**).

#### Association of epigenetic risk score with alcohol consumption

We conducted association between ERS and alcohol consumption in FHS. Each drink of self-reported alcohol consumption per day was associated with a 0.25-units higher ERS (*p* < 0.0001) in FHS. Consistent positive associations were also observed in three independent cohorts, GENOA, HRS, and Rhineland Study (**Supplemental Table 2**). One drink of self-reported alcohol consumption per day was associated with a 0.32-unit higher ERS in GENOA (*p* < 0.0001), with a 0.26-unit higher ERS in HRS (*p* < 0.0001), and with a 0.01-unit higher ERS in the Rhineland Study (*p* = 0.02). No significant association was observed in SHIP (**Supplemental Table 2**).

#### Association of alcohol consumption with blood pressure traits

Alcohol consumption was associated with SBP in both cross-sectional and longitudinal analyses in the FHS (**Supplemental Tables 12-13**). One additional drink/day alcohol consumption was associated with a 0.88 mm Hg higher SBP (*p* = 3.7e-4) in cross-sectional analysis and with a 0.92 mm Hg increase in SBP (*p* = 2.6e-4) between two exams in longitudinal analysis. We also found that one additional drink/day alcohol consumption was associated with 1.13 times of odds of being hypertension (95% CI = 1.04, 1.22; *p* = 0.006) in cross-sectional analysis. Associations of alcohol consumption with DBP were not significant (*p* > 0.05 in both cross-sectional and longitudinal analyses) in FHS (**Supplemental Tables 12-13**). In Rhineland and SHIP, we observed that higher alcohol consumption was significantly associated with SBP (Rhineland: β = 0.38, *p* = 0.006; SHIP: β = 2.94, *p* = 0.008) and DBP (Rhineland: β = 0.38, *p* = 1.6e-6; SHIP: β = 2.06, *p* = 0.002) (**Supplemental Table 12**). However, prevalent hypertension was not associated with alcohol consumption in either cohort. Whereas in HRS, higher level of DBP (β = 0.72, *p* = 0.04) and higher odds of hypertension (β = 0.17, *p* = 0.02) were associated with higher alcohol intake in cross-sectional analysis. No longitudinal significant association was observed between alcohol consumption and any of the blood pressure traits in GENOA or SHIP participants (**Supplemental Table 13**).

Additionally, we investigated whether the DNA methylation CpGs that were used for constructing the ERS were associated with blood pressure traits in cross-sectional analysis in FHS. When applying a relaxed threshold by false discovery rate (FDR) < 0.05, 26 and 17 of 144 CpG probes were significantly associated with SBP and DBP, respectively (**Supplemental Figures 6a-c, Supplemental Table 14-15**).

## DISCUSSION

This investigation builds upon our previous work that identified 144 CpGs that discriminated current heavy alcohol drinkers from non- and light-drinkers[15]. Using these CpGs, we constructed an alcohol consumption ERS that was associated with self-reported alcohol intake and explained additional variance (model R^2^ = 0.11 vs. 0.07 for AST and 0.14 vs. 0.11 for ALT) in hepatic biomarkers associated with alcohol intake. The risk score was associated with systolic and diastolic blood pressure in cross-sectional analyses both in discovery and replication analyses. In contrast, the risk score was not associated with longitudinal changes in blood pressure or incident hypertension either in discovery or replication studies.

The relationship between alcohol and hypertension is fairly well-defined with a comprehensive body of cross-sectional and longitudinal epidemiological studies, revealing a significant association between the two[34]. In addition, intervention studies and Mendelian randomization studies have suggested that the alcohol-blood pressure relationship is causal[7]. There are multiple mechanisms by which alcohol consumption can affect blood pressure. For example, chronic alcohol consumption has been reported to interfere with nitrous oxide (NO) production from endothelial cells, which affects various blood pressure regulatory mechanisms[35].

DNA methylation is an epigenetic mechanism that involves the transfer of a methyl group onto the fifth carbon position of the DNA building block, cytosine, to form 5-methylcytosine. This modification regulates gene expression by recruiting proteins to primarily inhibit the binding of transcription factor(s) to DNA. Therefore, certain cells may develop a stable and unique DNA methylation pattern that regulates tissue-specific gene expression. As previously mentioned, a prior study from our team[15] aimed to address the lack of reliable measures of alcohol intake by examining DNA methylation as a novel biomarker of alcohol use. As noted, DNA methylation provides a reliable measure of heavy alcohol intake and addresses a critical need.

While our analyses reveal that a risk score comprised of 144 CpGs reveals an association of current alcohol intake with blood pressure, it does not indicate an association of the risk score with longitudinal change in blood pressure. Several potential reasons may explain this finding. As follow-up examinations may be years apart for several of these participating cohorts, participants may change their lifestyle behaviors, which may affect methylation patterns. For example, participants may change their diet or initiate hypertensive medication during follow-up; participants who were moderate-to-heavy drinker may also reduce their alcohol consumption over time with aging. A similar finding was reported with longitudinal smoking traits and DNA methylation, where the majority of the differentially methylated CpG sites observed in analysis of current versus never smokers returned to the level of never-smokers within five years of smoking cessation[36]. The relationship between alcohol consumption and the degree of methylation changes over various periods of time is still being explored. A recent longitudinal study identified that 1414 CpGs were significantly associated with alcohol consumption in cross-sectional analysis while about a third of CpGs (n = 513) displayed associations between the changes in the methylation levels of these CpGs and the change of alcohol consumption between two exams[37]. This study indicated, as a dynamic measurement, DNA methylation may show stronger associations with cross-sectional behaviors and clinical phenotypes than associations with changes in traits during the follow-up, which was consistent with the findings in our study.

In addition, cross-sectional meta-analysis for SBP revealed significant heterogeneity. Our project performed a fixed-effects, inverse-variance weighted meta-analysis which assumes all independent cohorts are estimating the same underlying effect where the variation between estimates is attributed to random error. This heterogeneity may be attributable to a number of factors such as participants’ country or origin, ethnicity, age, sex, diet, and differences in sample collection methodology[38]. For example, the GENOA recruited participants who had at least 2 siblings diagnosed with essential hypertension before the 60-year-old[22]. Therefore, the GENOA participants may be more susceptible to hypertension than the general population due to common pathogenic genes and shared behavioral habits with their siblings. The SHIP recruited subjects from Northeast Germany, where the population had the lowest life expectancy in Germany at the time of recruitment[27]. These different recruitment criteria may distinguish participants in GENOA and SHIP from the general population, which may influence the relation between alcohol consumption, DNA methylation, and blood pressure traits. Furthermore, measurement error and a much higher SBP and DBP in GENOA and Rotterdam Study may partly explained differential associations between the ERS and alcohol consumption in these two studies, compared to results in other seven cohorts. Nevertheless, there was still a significant effect for this outcome, and our meta-analysis results support our findings in FHS.

Another key finding of our study was the improved model fit after the addition of the risk score in a cross-sectional, linear mixed regression model examining the relationship between self-reported alcohol intake and liver enzymes, ALT and AST, in FHS. While the association between self-reported alcohol intake and these liver enzymes was significant, the risk score was able to capture additional interindividual variations of the elevated liver enzymes (**Supplemental Table 7**). Whether this observation is due to the risk score as a tool that more accurately gauge degrees of alcohol consumption as survey or because the risk score represents other biological or environmental factors warrants future analyses.

This study has several limitations. Most of the participants are of European ancestry, which may render the findings of our project not applicable to other racial/ethnic populations. Secondly, we validated the utility of the risk score through association analysis with two enzymes associated with liver function, ALT and AST. While elevated levels of these enzymes are useful, common clinical indicators of chronic alcohol consumption, the levels of these enzymes can be modified by other means. Many prescription drugs such as cholesterol-lowering agents, anti-tuberculosis drugs, and non-steroidal anti-inflammatory drugs including aspirin are known to cause mildly elevated AST and ALT[39] which were not adjusted for. These enzyme levels can also be altered due to conditions such as autoimmune hepatitis: because of lacking validated data, FHS participants with such conditions were not excluded from analysis but cases of such conditions are supposed to be very few in the FHS. In addition, at the time of this study, the FHS did not have other biomarkers available for validation analysis such as gamma-glutamyl transferase which is another enzyme marker for heavy alcohol usage[40]. Our study also has several strengths. Our analyses showed that the risk score was significantly associated with self-reported alcohol intake data and with clinically-useful biomarkers of alcohol consumption. Our findings regarding the relationship between the risk score and blood pressure traits in FHS were replicated in meta-analysis of eight independent external cohorts.

## CONCLUSION

Our study developed an epigenetic risk score of alcohol consumption based on 144 alcohol-associated CpG probes identified in a large meta-analysis. This score showed an association with cross-sectional, but not longitudinal, blood pressure and prevalent hypertension among middle-aged and older participants. Our findings also provide a proof-of-concept that a DNA methylation-based epigenetic score capturing alcohol consumption can provide insights into alcohol-related disease risks, and further help to assess specific lifestyle factors that contribute to an individual’s health profile. This is especially pertinent in situations where self-reported behavioral data (e.g., alcohol consumption) are unavailable, susceptible to recall bias, or subject to significant data loss.

## Abbreviations

ERS: epigenetic risk score
SBP: systolic blood pressure
DBP: diastolic blood pressure
HTN: hypertension
AST: aspartate amino transferase
ALT: alanine aminotransferase

## AVAILABILITY OF DATA AND MATERIALS

Data and analytical codes that support our findings are available from the corresponding authors upon request.

## Data Availability

All data produced in the present study are available upon reasonable request to the authors.

## ACKNOWLEDGEMENTS

We thank all participants in the cohorts for their dedication in research.

## ETHICS DECLARATIONS

### Ethics approval and consent to participate

Institutional review committees of all cohorts approved this study. All study participants provided written informed consent. The FHS was reviewed and approved by The Boston Medical Center Institutional Review Boards. The ALHS was approved by the Institutional Review Board at National Institute of Environmental Health Sciences. The KORA cohort ethical approval was granted by the ethics committee of the Bavarian Medical Association (REC reference numbers: F4: #06068, FF4: #06068) and all were carried out in accordance with the principles of the Declaration of Helsinki. The GENOA was approved by Institutional Review Boards at the University of Michigan, University of Mississippi Medical Center, and Mayo Clinic. The HRS was approved by the University of Michigan. The MESA was approved by the Institutional Review Board of each study site, and written informed consent was obtained from all participants (IRB: #00009029). The Rhineland Study was approved by the ethics committee of the University of Bonn, Medical Faculty. The Rotterdam Study was approved by the medical ethics committee of the Erasmus University Medical Center, Rotterdam, the Netherlands. The SHIP was approved by the medical ethics committee of the University of Greifswald.

### Consent for publication

Not applicable.

## Contributions

D Levy, JM, and CL designed the study. HB, AK, M Wang, ML, SMR, LL, KSB, JDF, AP, CG, TD, SLRK, WZ, XG, JY, JIR, YL, XL, D Liu, JFT, GP, MMBB, IK, COR, TV, MG, JBJvM, MKN, MD, HJG, SJL, AT, M Waldenberger, DRW, JAS, D Levy, JM, and CL were involved in the data collection, data preparation, and statistical analyses. HB, AK, and M Wang drafted the manuscript. D Levy, JM, and CL supervised the study and provided critical review. All authors read and approved this manuscript.

