## Supplemental Material for "Association analysis between an epigenetic alcohol risk score and blood pressure"

### Supplemental Information

### Agricultural Lung Health Study

*Study description and history*

The Agricultural Lung Health Study (ALHS) is a case-control study of current asthma nested within the Agricultural Health Study (AHS) that enrolled farmers [licensed pesticide applicators] and their spouses from Iowa or North Carolina. There were 3301 participants enrolled in the ALHS in 2009-2013 (data version P3REL201209.00). The study was approved by the Institutional Review Board at National Institute of Environmental Health Sciences. Written informed consent was obtained from all participants.

*DNA methylation collection, processing, and additional analytical details*

Procedures for DNA methylation measurement and quality control have been outlined previously.^1^ In summary, DNA from whole blood was bisulfite converted using the EZ-96 DNA Methylation kit (Zymo Research Corporation) and assessed by Illumina’s MethylationEPIC BeadChip (EPIC array; Illumina Inc.). Samples with >5% of CpG with detection p > 1.0 x 10^-10^, intensity values less than three standard deviations below the mean bisulfite control intensities, or samples with sex mismatches were excluded from final analyses. Batch effect was corrected for using ComBat from the *sva* R package.^2^ Extreme methylation outliers were replaced with winsorized values using ewaff.handle outliers from the *ewaff* R package.^3^ After quality control, a total of 2,288 participants had available DNA methylation data. Cell type proportions (i.e., monocytes, neutrophils, eosinophils, natural killer cells, B cells, etc.) were estimated using the Houseman method^4^ with the Reinius reference panel.^5^

*Clinical and behavioral phenotype collection and measurement*

Field technicians collected blood samples and computer-assisted telephone interviews collected information on smoking history, current smoking status (yes/no), and alcohol consumption. For the collection of alcohol consumption, study participants answered “yes” or “no” to the question: “In the past 24 hours, have you had anything to drink that contained alcohol, including wine, wine coolers beer, or liquor?”. The participants also answered that questions “the amount, and types of alcoholic drinks consumed in the past 24 hours”. The final study sample included 1,293 individuals.

*Acknowledgements, funding, and disclosures*

The Agricultural Lung Health Study is supported by the Intramural Research Program of the National Institutes of Health, the National Institute of Environmental Health Sciences (Z01-ES102385, Z01-ES049030, Z01-ES043012) and the National Cancer Institute (Z01-CP010119). This work was also supported in part by American Recovery and Reinvestment Act funds. The authors declare no conflicts of interest.

### Cooperative Health Research in the Region Augsburg (KORA)

*Study description and history*

The Cooperative Health Research in the Region Augsburg (KORA) research platform has been collecting clinical and genetic data from the general population in the region of Augsburg, Germany for over 20 years. F4 (2006-2008) and FF4 (2013-2014) cohorts are follow-up studies from the KORA S4 (n=4,261) survey carried out 1999-2000. In the baseline examinations all inhabitants of German nationality between the ages of 25 and 74 years were enrolled. Participants completed a lifestyle questionnaire, including details on health status and medication use, underwent standardized examinations with blood samples taken.^6^

*DNA methylation collection, processing, and additional analytical details*

Genome-wide DNA methylation measurement at 485,577 genomic sites was performed using the Infinium HumanMethylation450K BeadChip (Illumina, Inc., CA, USA)^7^ in 1,802 KORA F4 samples. The laboratory process has been described previously.^8^ Briefly, denatured single-stranded genomic DNA was subjected to bisulfite treatment using the EZ-96 DNA Methylation Kit (Zymo Research, Orange, CA, USA). Bisulfite-converted samples were subjected to whole genome amplification, followed by enzymatic fragmentation and application to the BeadChips. The arrays were fluorescently stained and scanned with the Illumina HiScan SQ scanner. As readout, a methylated and an unmethylated signal count per CpG site are obtained. Counts were combined to β-values, defined as the ratio of the methylated signal intensity divided by the overall signal intensity: β-value, M/(M+U+α), where an offset was added as a regularization for the situation when both M and U are low, as recommended by Illumina.

DNA methylation data were preprocessed as follows. First, 65 probes that represent SNPs were excluded. Second, background correction was performed using the R package *minfi*, version 1.6.0.^9^ Third, detection p-values were defined as the probability of a signal being detected above the background signal level, as estimated from negative control probes. Consequently, signals with detection p-values ≥ 0.01 were removed, since they indicate putatively unreliable signals. Similarly, signals summarized from less than three functional beads on the chip were characterized as potentially unreliable and removed from the data set. Observations with less than 95% CpG sites providing reliable signals (75 in KORA F4) were excluded.

To reduce the non-biological variability between observations, data were normalized using quantile normalization on the raw signal intensities.^10^ Precisely, QN was stratified to six probe categories based on probe type and color channel (i.e., Infinium I signals from beads targeting methylated CpG sites obtained through the red and the green color channels, Infinium I signals from beads targeting unmethylated CpG sites obtained through the red and the green color channels, and Infinium II signals obtained through the red and the green color channels, as previously mentioned^7^) using the R package *limma*, version 3.16.5.^11^

*Clinical phenotype collection and measurement*

Unlike other cohorts, AF was not utilized as an exclusion criterion as KORA did not have this variable at the moment analysis was performed; however, prevalence of AF is low in this cohort regardless. With regard to smoking status, KORA participants were categorized into one of three groups: 1) the participant is either a regular smoker or a non-regular smoker (without specifying the number of cigarettes smoked), 2) the participant is a former smoker, and 3) the participants has never smoked. In the standardized interview, participant answered questions if they consumed alcohol in the past week or weekend. For those who answered yes, alcohol intake was assessed by the number of drinks of each types of alcoholic drinks over the weekend and on workdays in the previous week.^12^ Individuals at the time of DNA methylation measurement (KORA F4) were removed if they had previously had a myocardial infarct stationary treated, a stroke stationary treated, Angina pectoris (without regard to the exact localization), a history of coronary heart disease (in particular, one treated within the 12 months previous to DNA methylation measurement), a history of heart insufficiency (in particular, one treated within the 12 months previous to DNA methylation measurement). Individuals missing information regarding the above criteria or who responded “don’t know” were retained in the regression analysis. When a participant was missing DNA methylation value, since the number of missing CpGs was low, we set missing CpG value to 0, and thus, no individuals were excluded based on missing methylation data.

Finally, the regression models run were not adjusted for technical covariates as the ERS (LASSO) weights provided were determined based on residualized DNA methylation data already adjusted for technical covariates. A "pedigree" variable (genetic PCs) in the models was not used since in the KORA cohorts all participants are German. A “time between baseline and follow-up visit” variable was as well not used, because for each participant this period is actually very similar.

*Acknowledgements, funding, and disclosures*

The KORA cohort ethical approval was granted by the ethics committee of the Bavarian Medical Association (REC reference numbers: F4: #06068, FF4: #06068) and all were carried out in accordance with the principles of the Declaration of Helsinki. This covers consent for the use of biological material, including genetics. All research participants have signed informed consent prior to taking part in any research activities. The KORA data protection procedures were approved by the responsible data protection officer of the Helmholtz Zentrum München. The KORA study was initiated and financed by the Helmholtz Zentrum München –German Research Center for Environmental Health, which is funded by the German Federal Ministry of Education and Research (BMBF) and by the State of Bavaria. Furthermore, KORA research was supported within the Munich Center of Health Sciences (MC-Health), Ludwig-Maximilians-Universität, as part of LMUinnovativ.

**Framingham Heart Study (FHS)**

*Study description and history*

The FHS is a community-based, longitudinal cohort study initiated in 1948 to investigate CVD risk factors. In our development of the ERS, we utilized methylation data for participants in the FHS Offspring cohort who attended examination 8 (2005-2008)^13^ and Third Generation cohort participants who attended examination 2 (2008-2011).^14^ Since recruitment, both cohorts have been invited to attend research clinic health examinations every four to eight years to collect information on demographic and socioeconomic variables alongside clinical and laboratory data for CVD risk factors and outcomes.

*DNA methylation collection, processing, and additional analytical details*

Whole blood samples were assayed for DNA methylation via the Infinium Human Methylation 450 BeadChip platform (San Diego, CA). The methylated probe intensity and total probe intensities were extracted using the Illumina Genome Studio (version 2011.1) with the methylation module (version 1.9.0). Preprocessing of the methylated (M) signal and unmethylated signal (U) was conducted; methylation beta-value (βM) was defined as β=M/(U+M). FHS used the function “DASEN” of the *wateRmelon* R package (version 3.0.2)^15^ to normalize the methylation β proportion. Further information regarding DNA extraction and processing have been outlined.^16^

*Clinical phenotype collection and measurement*

Participant weight was measured with the participant in a gown and without shoes, and a calibrated scale was used and weight was rounded to the nearest pound. Participant height was measured using a vertically-mounted stadiometer and rounded down to the neatest ¼ inch. Body mass index (BMI) was calculated according to the formula: $\frac{Weight (lbs)}{Height {(in)}^{2}}\times703$. Cigarette smoking status was defined based on categorical self-reported smoking behavior. Current smokers were defined as participants who have smoked on average at least one cigarette per day in the past year; former smokers were defined as participants who have previously smoked on average at least one cigarette per day but has stopped smoking for at least one year; never smokers were defined as participants who never smoked. The collection of alcohol consumption was previous detailed. In brief, alcohol consumption data was collected with a stand-alone questionnaire that includes specific questions to collect usual frequency and types of alcoholic beverage (i.e., beer, wine, or liquor), on average, consumed by a participant per week over the past year at each health examination.^17^

*Acknowledgements, funding, and disclosures*

The Framingham Heart Study was supported by NIH contract N01-HC-25195. The analytical component of this project was funded by the Division of Intramural Research, National Heart, Lung, and Blood Institute, National Institutes of Health, Bethesda, MD (D. Levy, Principal Investigator). J. Ma is supported by NIH grant R01AA028263; C. Liu is supported by R01AA028263 and R01HL15569 grants. The views and opinions expressed in this manuscript are those of the authors and do not necessarily represent the views of the National Heart, Lung, and Blood Institute, the National Institutes of Health, or the U.S. Department of Health and Human Services. All authors declare no conflicts of interest.

**Genetic Epidemiology Network of Arteriopathy (GENOA)**

*Study description and history*

The Genetic Epidemiology Network of Arteriopathy (GENOA) study is a community-based study of hypertensive sibships that was designed to investigate the genetics of hypertension and target organ damage in African Americans from Jackson, Mississippi and non-Hispanic whites from Rochester, Minnesota.^18^ In the initial phase of GENOA (Phase I: 1996-2001), all members of sibships containing ≥ 2 individuals with essential hypertension clinically diagnosed before age 60 were invited to participate, including both hypertensive and normotensive siblings. Exclusion criteria of the GENOA study were secondary hypertension, alcoholism or drug abuse, pregnancy, insulin-dependent diabetes mellitus, or active malignancy. Eighty percent of African Americans (1,482 participants) and 75% of non-Hispanic whites (1,213 participants) from the initial study population returned for the second examination (Phase II: 2001-2005). Study visits were made in the morning after an overnight fast of at least eight hours. Demographic information, medical history, clinical characteristics, lifestyle factors, and blood samples were collected in each phase. Written informed consent was obtained from all participants and approval was granted by participating institutional review boards. DNA methylation levels were measured only in African American participants.

*DNA methylation collection, processing, and additional analytical details*

A total of 1,106 samples at Phase I and 304 samples at Phase II were assessed using the Illumina HumanMethylationEPIC BeadChip. First, raw IDAT files were imported using *minfi* R package.^9^ We used the *shinyMethyl* R package^19^ to visualize the raw intensity data and identify sex mismatches and outliers, which were removed. We also obtained detection p-value for each sample at each probe, and individual probes with detection p-value <10-16 were considered to be detected successfully.^10^ Samples and probes with detection rate <10% were removed. Samples with incomplete bisulfite conversion identified using the “QCinfo” function in the *ENmix* R package were removed.^20^ We also checked sample identity using the 59 SNP probes implemented in the EPIC chip and removed mismatched samples. Next, Noob was used for individual background and dye-bias normalization.^21^ Since two types of probes are present on the EPIC BeadChip (Infinium I and Infinium II), we used the Regression on Correlated Probes (RCP) method to adjust for probe-type bias.^22^ After exclusions, a total of 857,121 probes in 1,100 samples at Phase I and 294 samples at Phase II were available for analysis.

In cross-sectional and prospective analyses, familial relationships within GENOA were taken into account by including a random effect for family ID in GEE models used to assess relationships between the ERS and blood pressure traits. The SAS (v9.4 TS1M2) procedure GENMOD was used for this purpose. Blood pressures were analyzed as normally distributed linear traits, and hypertension was analyzed as binomially distributed with logit link.

*Clinical phenotype collection and measurement*

The study sample consisted of 1,053 participants with methylation data and no self-reported history of heart attack, myocardial infarction, or heart surgery at Phase I. For follow-up longitudinal models, 1,037 participants had blood pressure data at Phase II. Excluding those with hypertension at Phase I, a total of 325 were used for incident hypertension analysis. All covariates were taken from Phase I.

Blood pressure measures were taken as the average of the last two (out of 3) measurements taken during the exam. Systolic and diastolic averages were adjusted by 15 mmHg and 10 mmHg, respectively, for those on antihypertensive medication. Smoking status was categorized as never (smoked fewer than 100 cigarettes in lifetime), former (smoked 100 cigarettes but do not currently smoke), and current (currently smoke). Height (cm) and weight (kg) were measured at in-person exams and converted to inches and pounds, respectively. Alcohol consumption was calculated as the number of drinks per week based on aggregated measurements of a variety of alcoholic drinks.^23^

*Acknowledgements, funding, and disclosures*

Support for the Genetic Epidemiology Network of Arteriopathy (GENOA) was provided by the National Heart, Lung and Blood Institute (U10 HL054457, RC1 HL100185, R01 HL087660, R01 HL119443, R01 HL141292, and R01 HL133221). We would also like to thank the families that participated in the GENOA study.

**Health and Retirement Study (HRS)**

*Study description and history*

The Health and Retirement Study (HRS) is a longitudinal survey of a representative sample of Americans over the age of 50.^24^ Over 42,000 persons in 26,000 households have been interviewed since 1992. The study interviews respondents every two years about income and wealth, health and use of health services, work and retirement, and family connections. The sample is refreshed periodically with a new cohort of respondents to offset attrition and death and maintain ~20,000 individuals per sample wave. Starting in 2006, half of the core sample is randomly assigned to a face-to-face interview enhanced with physical and biological measures and a mail-back psychosocial questionnaire. The other half is interviewed by telephone. Each interview mode then alternates between the two half-samples in subsequent waves.

*DNA methylation collection, processing, and additional analytical details*

As part of the Venous Blood Study (VBS), an ancillary study of HRS, venous blood was collected in the 2016 wave. For a total of 4,103 respondents, DNA methylation was assessed from the blood samples using the Illumina HumanMethylationEPIC BeadChip. Data were imported using the *minfi* R package.^9^ Sex mismatches and with an average median intensity <8.5 were removed. A detection p-value<0.01 was used to remove samples and probes with detection rate <5%. Sample identity was assessed using 59 SNP probes on the EPIC chip, but no additional samples were removed. After exclusions, a total of 836,660 probes in 4,018 samples were available for analysis.

With regards to cross-sectional and prospective analyses between the ERS and blood pressure traits, blood pressures were analyzed as normally distributed linear traits with the SAS (v9.4 TS1M2) procedure GENMOD, and hypertension was analyzed as binomially distributed with logit link using the SAS procedure LOGISTIC.

*Clinical phenotype collection and measurement*

The study samples consisted of 1348 participants from the 2016 wave (concurrent with methylation) and 1196 participants from the 2018 wave that had methylation data and no self-reported history of heart attack, coronary heart disease, angina, congestive heart failure, or other heart problems at the 2016 wave. Blood pressure measures were taken as the average of three measurements taken during the face-to-face interview, and both systolic and diastolic averages were adjusted by 15 mmHg and 10 mmHg, respectively, for those on antihypertensive medication. Smoking status was categorized as never, former, and current. At each wave, participants were asked if they ever drank alcohol. For the ones who answered yes, they were asked, on average, how many days and the number drinks of alcohol they consumed in the last three months.^25^ Height (in) and weight (lbs) were measured at face-to-face examinations.

*Acknowledgements, funding, and disclosures*

HRS is supported by the National Institute on Aging (NIA U01AG009740). The HRS DNA methylation was measured by the University of Minnesota Genomics Center (UMGC). Sample coordination, plate design, and quality control was performed by the University of Minnesota Advanced Research and Diagnostics Laboratory (ARDL).

**Multi-Ethnic Study of Atherosclerosis (MESA)**

*Study description and history*

The Multi-Ethnic Study of Atherosclerosis (MESA) is a study of the characteristics of subclinical cardiovascular disease and the risk factors that predict progression to clinically overt cardiovascular disease or progression of the subclinical disease. MESA consisted of a diverse, population-based sample of an initial 6,814 asymptomatic men and women aged 45-84. 38 percent of the recruited participants were white, 28 percent African American, 22 percent Hispanic, and 12 percent Asian, predominantly of Chinese descent. Participants were recruited from six field centers across the United States: Wake Forest University, Columbia University, Johns Hopkins University, University of Minnesota, Northwestern University and University of California - Los Angeles. This study was approved by the IRB of each study site, and written informed consent was obtained from all participants (IRB: #00009029). Each participant received an extensive physical exam and determination of coronary calcification, ventricular mass and function, flow-mediated endothelial vasodilation, carotid intimal-medial wall thickness and presence of echogenic lucencies in the carotid artery, lower extremity vascular insufficiency, arterial wave forms, electrocardiographic (ECG) measures, standard coronary risk factors, sociodemographic factors, lifestyle factors, and psychosocial factors. Selected repetition of subclinical disease measures and risk factors at follow-up visits allowed study of the progression of disease. Participants are being followed for identification and characterization of cardiovascular disease events, including acute myocardial infarction and other forms of coronary heart disease (CHD), stroke, and congestive heart failure; for cardiovascular disease interventions; and for mortality. The first examination took place over two years, from July 2000 - July 2002. It was followed by five examination periods that were 17-20 months in length, including the latest completed exam 6 (2016-2018) with high cohort retention.

*DNA methylation collection, processing, and additional analytical details*

Microarray technology was used to assay methylation at CpG sites in genomic DNA samples. The assays were performed using the Illumina EPIC array, which targets over 850,000 CpG sites. DNA samples were treated with bisulfite to measure 5-methylcytosine (5-mC) and 5-hydroxymethylcytosine (5-hmC): ~200 of these matched samples were also treated with an oxidizing reagent to distinguish between 5-mC and 5-hmC. Beta values will be corrected for background and normalized by standard procedures (e.g., BMIQ, SWAN, DASEN). Quality control procedures will include removing sites with detection p-values p > 0.05 in more than 5-20% of samples and removing samples with detection p-values p > 0.05 in > 5% of sites.

 DNAm was estimated as the proportion of methylated beads relative to combined unmethylated and methylated beads for a specific CpG site defined as the β value (ranging from 0 [unmethylated] to 1 [methylated]). All methylation data were normalized using beta-mixture quantile normalization. Technical covariates included chip and row to adjust for batch effects and cell composition, which was estimated using the reference-based Houseman method.^4^

*Clinical phenotype collection and measurement*

The study sample consisted of 935 participants with methylation data and no self-reported history of CHD at Exam1. For follow-up longitudinal models, 935 of these had blood pressure data at exam 5. Excluding those with hypertension at Exam1, 566 (203 incident cases) were left for incident hypertension analysis. All covariates were taken from Exam1.

Systolic and diastolic BP (SBP, DBP) were measured at each exam following a standard protocol. Participants rested for 5 minutes and then three measurements were taken at 2-minute intervals using an automated oscillometric sphygmomanometer. The second and third measurements were averaged and this value was used for analysis. To account for treatment effects of antihypertensive medication use, we added 15 mmHg to the observed SBP and 10 mmHg to the observed DBP values among participants reporting antihypertensive medication use. Smoking status was categorized as never, former, and current. Height (cm) and weight (lbs) were measured at in-person exams. The information on the collection of alcohol consumption was previously collected.^26^ In brief, alcohol consumption was collected by trained personnel using personal‐history questionnaire. The participants were asked the following questions: “Have you ever consumed alcoholic beverages?” If yes, the following question was, “Do you presently drink alcoholic beverages?” For those who reported former drinking were asked about the usual number of drinks consumed per week before they stopped drinking. Those who reported current drinking also were asked about the number of drinks consumed during the past 24 hours and the largest number of drinks consumed in 1 day in the past month.

*Acknowledgements, funding, and disclosures*

Whole genome sequencing (WGS) for the Trans-Omics in Precision Medicine (TOPMed) program was supported by the National Heart, Lung and Blood Institute (NHLBI). WGS for “NHLBI TOPMed: Multi-Ethnic Study of Atherosclerosis (MESA)” (phs001416.v1.p1) was performed at the Broad Institute of MIT and Harvard (3U54HG003067-13S1). Centralized read mapping and genotype calling, along with variant quality metrics and filtering were provided by the TOPMed Informatics Research Center (3R01HL-117626-02S1). Phenotype harmonization, data management, sample-identity QC, and general study coordination, were provided by the TOPMed Data Coordinating Center (3R01HL-120393-02S1), and TOPMed MESA Multi-Omics (HHSN2682015000031/HSN26800004). The MESA projects are conducted and supported by the National Heart, Lung, and Blood Institute (NHLBI) in collaboration with MESA investigators. Support for MESA is provided by contracts 75N92020D00001, HHSN268201500003I, N01-HC-95159, 75N92020D00005, N01-HC-95160, 75N92020D00002, N01-HC-95161, 75N92020D00003, N01-HC-95162, 75N92020D00006, N01-HC-95163, 75N92020D00004, N01-HC-95164, 75N92020D00007, N01-HC-95165, N01-HC-95166, N01-HC-95167, N01-HC-95168, N01-HC-95169, UL1-TR-000040, UL1-TR-001079, UL1-TR-001420, UL1TR001881, DK063491, and R01HL105756. The authors thank the other investigators, the staff, and the participants of the MESA study for their valuable contributions. A fill list of participating MESA investigators and institutes can be found at [http://www.mesa-nhlbi.org](https://nam02.safelinks.protection.outlook.com/?url=http%3A%2F%2Fwww.mesa-nhlbi.org%2F&data=04%7C01%7Cwpost%40jhmi.edu%7Ce250d1c265a847ec090f08d9fbc439a1%7C9fa4f438b1e6473b803f86f8aedf0dec%7C0%7C0%7C637817642593854966%7CUnknown%7CTWFpbGZsb3d8eyJWIjoiMC4wLjAwMDAiLCJQIjoiV2luMzIiLCJBTiI6Ik1haWwiLCJXVCI6Mn0%3D%7C3000&sdata=tVqOVyqCZnioV87T5M39EUUQjtfxVyF4i%2FhQtZTLbwY%3D&reserved=0). This study also supported in part by NIH contract R01HL155569.

**Rhineland Study**

*Study description and history*

The Rhineland Study is an ongoing single-center, population-based cohort study that recruits people aged 30 years and above from two geographically defined areas in Bonn, Germany. The only exclusion criterion is insufficient command of the German language to provide informed consent. Persons living in the recruitment areas are predominantly German from Caucasian descent. A primary objective of the Rhineland Study is to identify determinants and markers of healthy aging with a deep-phenotyping approach. At baseline, participants complete an 8-hour in-depth multi-domain phenotypic assessment and various types of biomaterials (blood, urine, stool, and hair) are collected. Approval to undertake the study was obtained from the ethics committee of the University of Bonn, Medical Faculty. We obtained written informed consent from all participants in accordance with the Declaration of Helsinki.

*DNA methylation collection, processing, and additional analytical details*

Genomic DNA was extracted from buffy coat fractions of anti-coagulated blood samples using Chemagic DNA buffy coat kit (PerkinElmer, Germany), and was subsequently bisulfite converted using the DNA methylation kit according to the manufacturer’s instructions. DNA methylation levels were measured using Illumina’s Human MethylationEPIC BeadChip. The methylation level for each probe was derived as a beta value representing the fractional level of DNA methylation at that probe. Sample-level and probe-level quality control was performed using the *minfi* package in R (version 3.5.0).^21^ Samples with sex mismatch or a missing rate at > 1% across all probes were excluded. Probes with a missing rate > 1% (at a detection p-value > 0.01) were also excluded following previously published recommendation guidelines for analyzing methylation data.^27^

*Clinical phenotype collection and measurement*

Alcohol intake was assessed using a self-administered semi-quantitative food frequency questionnaire (FFQ). The questionnaire was originally developed for the European Prospective Investigation into Cancer and Nutrition (EPIC) study in Potsdam^28^ and adapted for the Rhineland Study. Participants were queried about their regular consumption of food and beverages, including various alcoholic drinks such as beer, wine, liqueurs, and spirits, over the past 12 months. For each alcoholic beverage, respondents had seven frequency options to choose from, ranging from “never” to “3 times per day or more.” Additionally, participants were asked to indicate the typical number of standard portion sizes they consumed for each type of alcoholic beverage, with five options available (1/2, 1, 2, 3, 4, or more). Daily total alcohol intake in g/day for each participant was calculated as the sum of alcohol content across all beverages consumed.^28^ Systolic blood pressure (SBP, mmHg) and diastolic blood pressure (DBP, mmHg) were measured three times (separated by ten-minute intervals), using an oscillometric blood pressure device (Omron 705 IT). The measurements were performed while people were sitting in a resting chair in a quiet environment, and the average of the second and third measurements was used for further calculation. In this analysis, to account for the treatment effects of antihypertensive medication use, we added 15 mmHg to the measured SBP and 10 mmHg to the measured DBP values among participants reporting antihypertensive medication use. Smoking status was defined as “current smoker” or “non-current smoker” based on self-report. Missing smoking values were imputed based on cotinine metabolite levels: individuals with a cotinine level exceeding the non-current smoker sample-defined 97.5 percentile were classified as current smokers.

For the current analyses, we used baseline data of the first 4,435 participants of the Rhineland Study with methylation data. We excluded one sample with ERS outlier, 16 samples with energy intake lower than 800 kcal/day or higher than 6000 kcal/day and 65 samples that had missing values in blood pressure traits. We further excluded participants with cardiovascular diseases (n = 386). The final analysis sample comprised 3,968 participants.

*Acknowledgements, funding, and disclosures*

The nutrition and omics analyses in the Rhineland Study were supported by the Diet-Body-Brain Competence Cluster in Nutrition Research funded by the Federal Ministry of Education and Research (grant numbers 01EA1410C and FKZ:01EA1809C). This work was further supported through SFB 1454– project number 432325352 by the Deutsche Forschungsgemeinschaft (DFG, German Research Foundation). The authors declare no conflicts of interest.

**Rotterdam Study**

*Study description and history*

The Rotterdam Study (RS) is a prospective population-based study started in 1989.^29^ It is composed of residents of the neighborhood of Ommoord, Rotterdam, the Netherlands, aged 40 years and over, who have been recruited in four sub-cohorts: RS-I, RS-II, RS-III and RS-IV. Each participant gave an informed consent and the study was approved by the medical ethics committee of the Erasmus University Medical Center, Rotterdam, the Netherlands.

*DNA methylation collection, processing, and additional analytical details*

DNA for methylation measurement was extracted from whole peripheral blood (stored in EDTA tubes) by standardized salting out methods, from a randomly selected sample of RS second (RSII) and third (RSIII) sub-cohorts. At the Genetic Laboratory (Department of Internal Medicine, Erasmus University Medical Center, Rotterdam, the Netherlands), genome-wide DNA-methylation levels in 1,613 subjects from the Rotterdam Study were determined using the Illumina HumanMethylation450K BeadChip arrays (Illumina, Inc., San Diego, CA, USA) according to the manufacturers protocol. Genomic DNA was first extracted from whole peripheral blood by standardized salting out methods. Samples were then bisulfite treated using the Zymo EZ-96 DNA-methylation kit (Zymo Research, Irvine, CA, USA), and subsequently hybridized to the arrays. Preparation and normalization of this array data was performed according to the CPACOR workflow using the software package R (www.r-project.org). The idat files were read using the *minfi* package. Samples showing inadequate hybridization, incomplete bisulfite treatment and gender swaps were excluded. Samples were excluded if signal detection rate across probes was < 95%. CpG methylation proportion was reported as a normalized β-value ranging from 0 to 1, where 1 represents 100% methylation. β values being outside 1.5 times the interquartile range per probe were set to missing. To avoid sex bias, CpGs annotated to genes in sex chromosomes were excluded. Methylation data for two out of the 144 CpGs were missing; therefore, the ERS was calculated using the remaining 142 CpGs only.

*Clinical phenotype collection and measurement*

For the analysis presented, we used data from 1,109 participants at baseline examination. Participants underwent interviews and health examinations from trained physicians. Alcohol intake data was captured by either dietary interview administered at home or food frequency questionnaire, depending on the follow up visit of the participants. Participants were asked specific question regarding type of alcoholic beverage consumed (i.e. red wine, white wine, beer and strong alcoholic beverages) and quantities of these types of beverages consumed in the past 24 hours and during a typical week.^30^ The information gathered was used to compute an average alcohol consumption in “grams per day.”

*Acknowledgments, funding, and disclosures*

The Rotterdam Study is supported by the Erasmus MC University Medical Center and Erasmus University Rotterdam; The Netherlands Organization for Scientific Research (NWO); The Netherlands Organization for Health Research and Development (ZonMw); the Research Institute for Diseases in the Elderly (RIDE); The Netherlands Genomics Initiative (NGI); the Ministry of Education, Culture and Science; the Ministry of Health, Welfare and Sports; the European Commission (DG XII); and the Municipality of Rotterdam. The contribution of inhabitants, general practitioners and pharmacists of the Ommoord district and management team to the Rotterdam Study is gratefully acknowledged.

**Study of Health in Pomerania (SHIP)**

*Study description and history*

The Study of Health in Pomerania (SHIP-Trend) is a longitudinal population-based cohort study in West Pomerania, a region in the northeast of Germany, assessing the prevalence and incidence of common population-relevant diseases and their risk factors. Baseline examinations for SHIP-Trend were carried out between 2008 and 2012, comprising 4,420 participants aged 20 to 81 years. Study design and sampling methods were previously described.^31^ The medical ethics committee of the University of Greifswald approved the study protocol, and oral and written informed consents were obtained from each of the study participants.

*DNA methylation collection, processing, and additional analytical details*

DNA was extracted from blood samples of 508 SHIP-Trend participants to assess DNA methylation using the Illumina HumanMethylationEPIC BeadChip array. Samples were randomly selected based on availability of multiple OMICS data, excluding type II diabetes, and enriched for prevalent MI. The samples were taken between 07:00 AM and 04:00 PM, and serum aliquots were prepared for immediate analysis and for storage at -80 °C in the Integrated Research Biobank (Liconic, Liechtenstein). Processing of the DNA samples was performed at the Helmholtz Zentrum München. Preparation and normalization of the array data was performed according to the CPACOR workflow^10^ using the software package R (www.r-project.org). The array idat files were processed using the *minfi* package. Probes that had a detection p-value above background (sum of per-array methylated and unmethylated intensity values based p-value ≥ 1E-16) were set to missing. Methylation beta values were calculated as proportion of methylated intensity value on the sum of methylated+unmethylated+100 intensities. Arrays with observed technical problems (± 4 standard deviations outside control probe intensity mean) during steps like bisulfite conversion, hybridization or extension, as well as arrays with mismatch between sex of the proband and sex determined by the chr X and Y probe intensities were removed from subsequent analyses. Additionally, only arrays with a call rate ≥ 95% were processed further resulting in 495 samples with methylation data on 865,859 sites available for subsequent analyses.

To account for potential confounding effects due to blood cell composition, blood cell subtypes were estimated by the Houseman method^4^ and included in the association model. Additionally, the array processing batch (n=248 and n=247) and the first six principal components of the control probe intensities obtained by the CPACOR workflow were included in the model to account for technical factors. Details on assessment of the metabolic phenotypes and covariates used in this analysis are provided within the SHIP cohort design.

*Clinical phenotype collection and measurement*

Alcohol consumption was assessed as described in a previous publication by Baumeister et al.^32,33^ Through questionnaire, participants were asked about the frequency and volume of their alcohol consumption over the last 30 days. Responses to the questions were expressed as absolute grams or alcohol per day with 12 grams of alcohol equivalent to approximately one bottle of beer, one glass of wine or one mixed drink.

*Acknowledgments, funding, and disclosures*

SHIP is part of the Community Medicine Research net of the University of Greifswald, Germany, which is funded by the Federal Ministry of Education and Research (grants no. 01ZZ9603, 01ZZ0103, and 01ZZ0403), the Ministry of Cultural Affairs as well as the Social Ministry of the Federal State of Mecklenburg-West Pomerania, and the network ‘Greifswald Approach to Individualized Medicine (GANI_MED)’ funded by the Federal Ministry of Education and Research (grant 03IS2061A). DNA methylation data have been supported by the DZHK (grant 81X3400104). The University of Greifswald is a member of the Caché Campus program of the InterSystems GmbH. HJG has received travel grants and speakers honoraria from Fresenius Medical Care, Neuraxpharm, Servier and Janssen Cilag as well as research funding from Fresenius Medical Care.

### Supplemental Tables (in a separate excel file)

### Supplemental Table 1. Clinical characteristics at baseline examination

**Supplemental Table 2.** Clinical characteristics at follow-up examination

**Supplemental Table 3**. ALHS self-reported alcohol intake

**Supplemental Table 4**. Summarized information of alcohol consumption and blood pressure traits across all participating cohorts

**Supplemental Table 5**. ERS distribution across all participating cohorts

**Supplemental Table 6**. Reduced vs. full model analysis with ERS and liver enzymes

**Supplemental Table 7**. Cross-sectional and longitudinal analysis results for hypertension with 130/80 mmHg in FHS

**Supplemental Table 8**. Cross-sectional analysis results for ERS and blood pressure traits

**Supplemental Table 9**. Prospective analysis results for ERS and blood pressure traits

**Supplemental Table 10**. Cross-sectional analysis results for ERS and blood pressure traits among untreated participants

**Supplemental Table 11**. Cross-sectional analysis results for ERS and alcohol consumption

**Supplemental Table 12**. Cross-sectional analysis results for alcohol consumption and blood pressure traits

**Supplemental Table 13**. Longitudinal analysis results for alcohol consumption and blood pressure traits

**Supplemental Table 14**. Cross-sectional analysis results for systolic blood pressure and 144 CpG probes in FHS

**Supplemental Table 15**. Cross-sectional analysis results for diastolic blood pressure and 144 CpG probes in FHS

**Supplemental Figures**

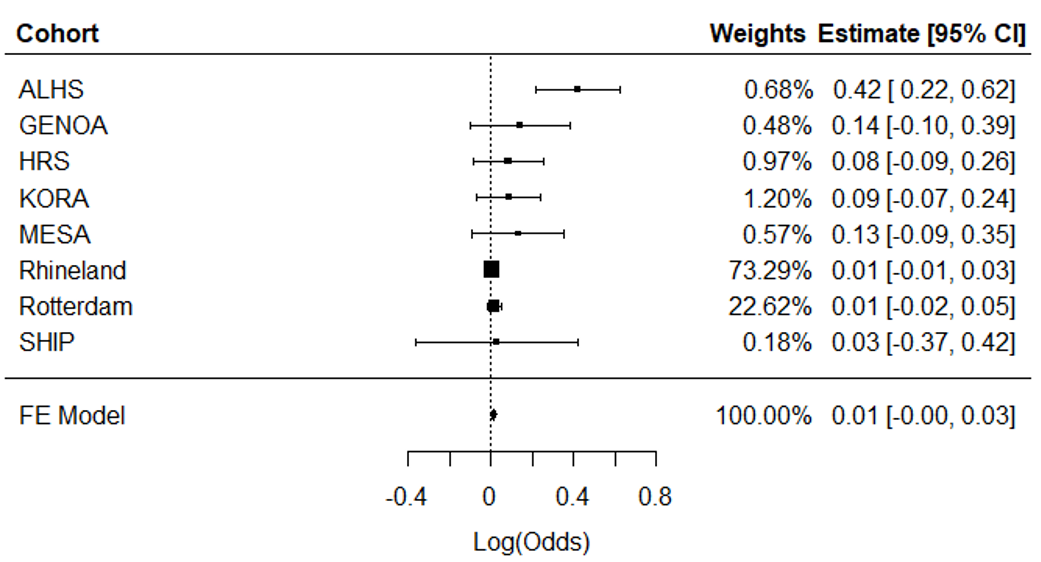

**Supplemental Figure 1.** Meta-analysis of cross-sectional association analyses of ERS in relation to hypertension in eight independent cohorts (n = 11,544). ALHS, Agricultural Lung Health Study; GENOA, Genetic Epidemiology Network of Arteriopathy; HRS, Health and Retirement Study; KORA, Cooperative Health Research in the Region Augsburg; MESA, Multi-Ethnic Study of Atherosclerosis; SHIP, Study of Health in Pomerania; FE, Fixed Effect; 95% CI, 95% Confidence Interval; log(Odds), log odds ratio.

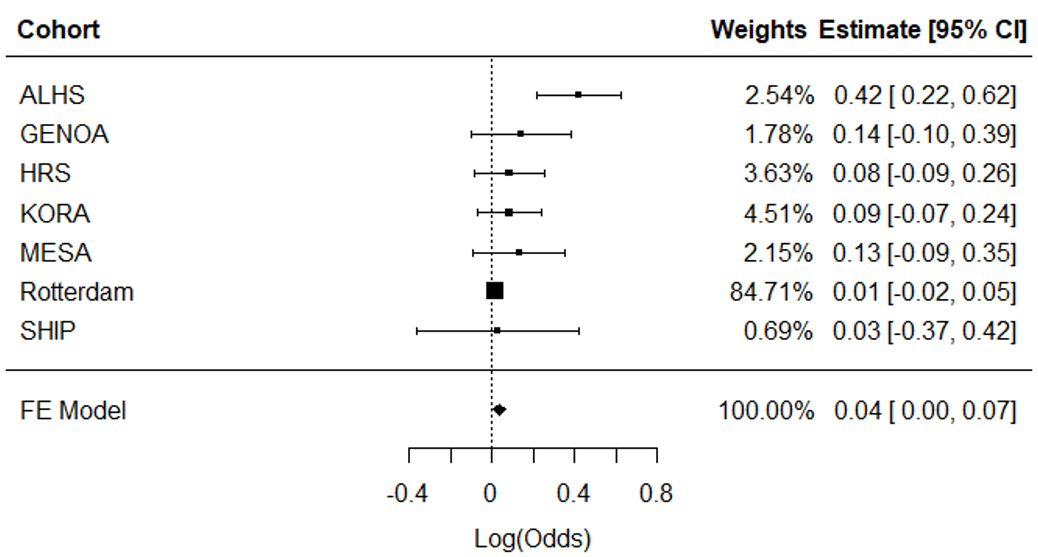

**Supplemental Figure 2.** Meta-analysis of cross-sectional association analyses of ERS in relation to HTN (excluding the Rhineland Study as sensitivity analysis). ALHS, Agricultural Lung Health Study; GENOA, Genetic Epidemiology Network of Arteriopathy; KORA, Cooperative Health Research in the Region Augsburg; HRS, Health and Retirement Study; MESA, Multi-Ethnic Study on Atherosclerosis; SHIP, Study of Health in Pomerania; FE, Fixed Effect; 95% CI, 95% Confidence Interval; log(Odds), log odds ratio.

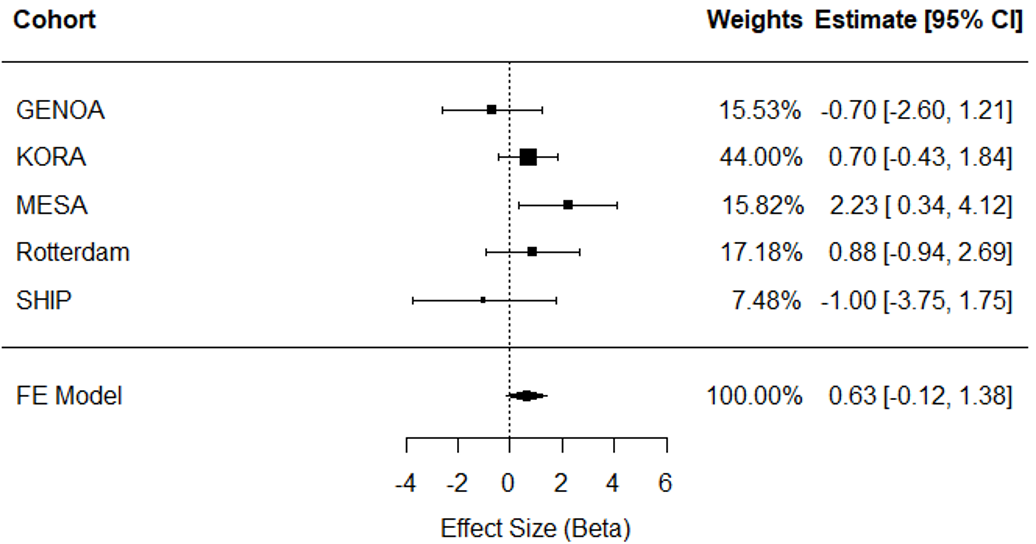

**Supplemental Figure 3.** Meta-analysis of longitudinal association analyses of ERS in relation to changes in systolic blood pressure in five independent cohorts (n=3,910). GENOA, Genetic Epidemiology Network of Arteriopathy; KORA, Cooperative Health Research in the Region Augsburg; MESA, Multi-Ethnic Study of Atherosclerosis; SHIP, Study of Health in Pomerania; FE, Fixed Effect; 95% CI, 95% Confidence Interval.

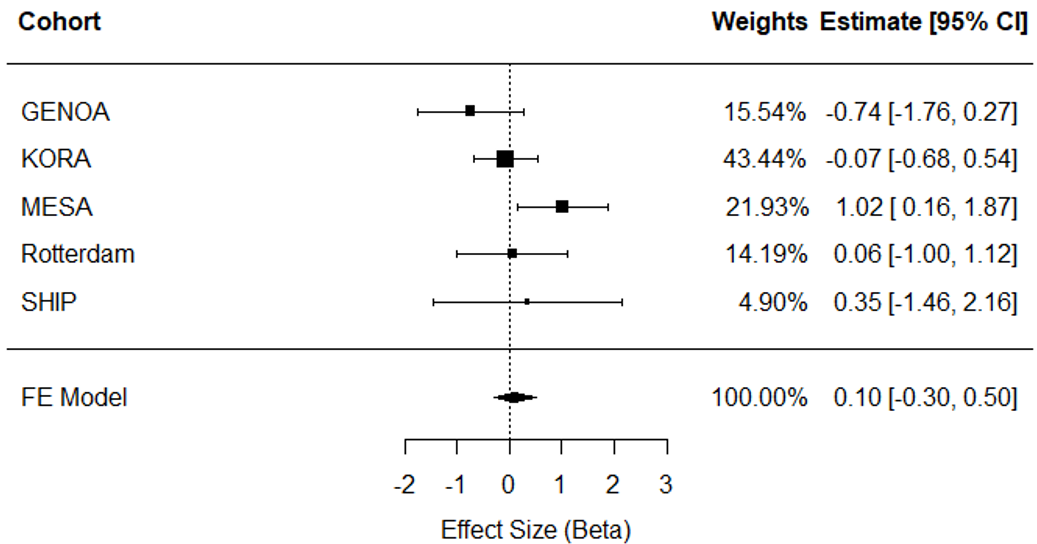

**Supplemental Figure 4.** Meta-analysis of longitudinal association analyses of ERS in relation to changes in diastolic blood pressure in five independent cohorts (n=3,910). GENOA, Genetic Epidemiology Network of Arteriopathy; KORA, Cooperative Health Research in the Region Augsburg; MESA, Multi-Ethnic Study of Atherosclerosis; SHIP, Study of Health in Pomerania; FE, Fixed Effect; 95% CI, 95% Confidence Interval.

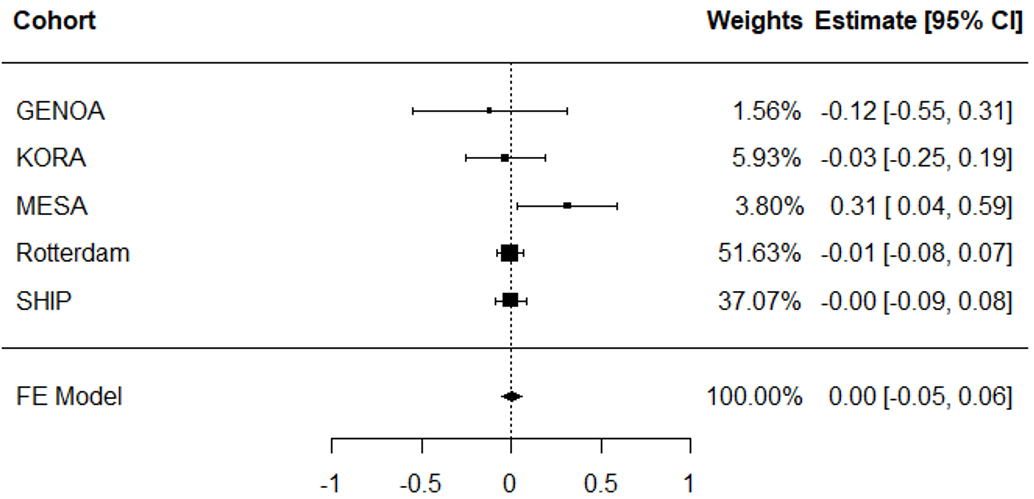

**Supplemental Figure 5.** Meta-analysis of ERS in relation to incident (i.e., new-onset) hypertension in five independent cohorts (n=3,228)**.** GENOA, Genetic Epidemiology Network of Arteriopathy; KORA, Cooperative Health Research in the Region Augsburg; MESA, Multi-Ethnic Study of Atherosclerosis; SHIP, Study of Health in Pomerania; FE, Fixed Effect; 95% CI, 95% Confidence Interval.

a. b.

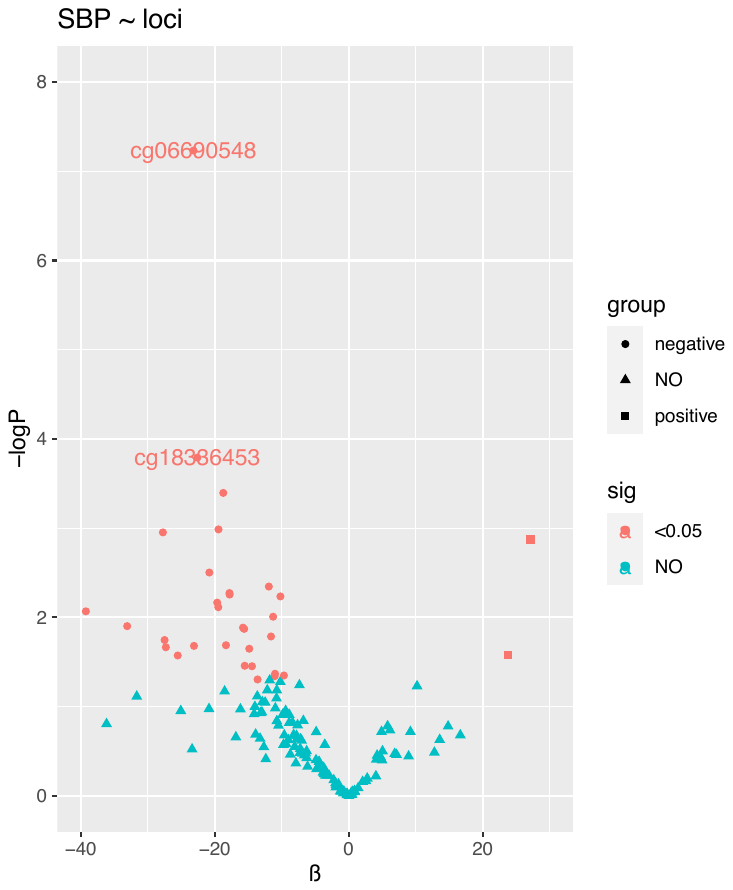

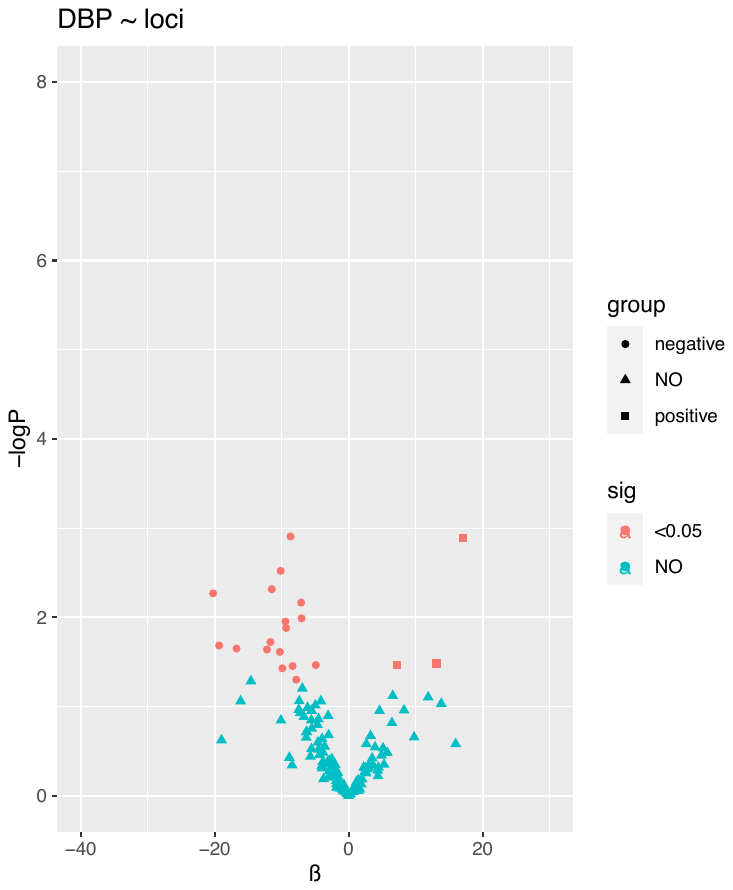

c.

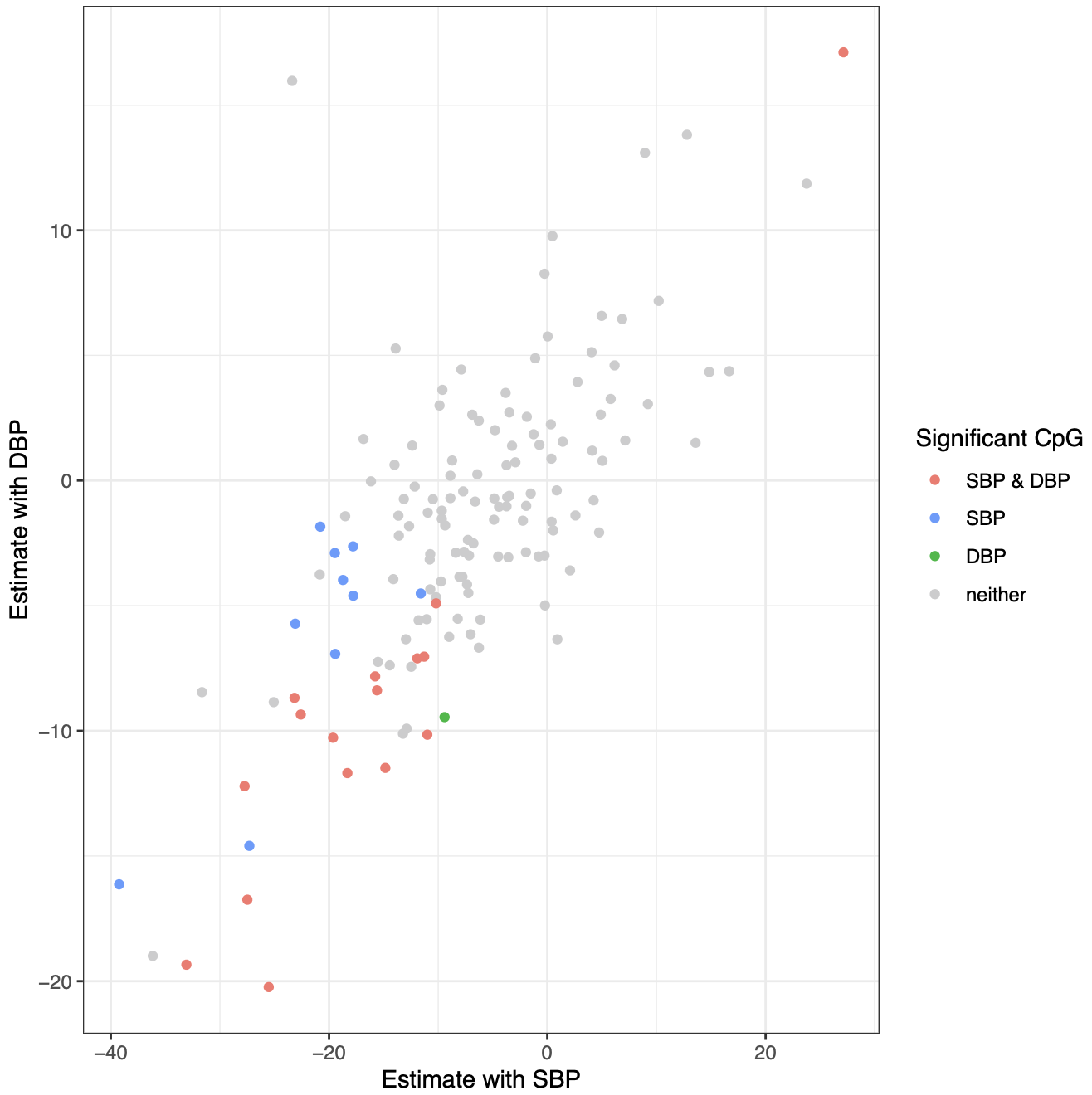

**Supplemental Figure 6** Cross-sectional analysis of 144 CpG probes with blood pressure traits (i.e., SBP and DBP) in FHS. a) 144 CpG probes with SBP; b) 144 CpG probes with DBP; c) comparison on estimates and p-value of 144 CpG probes between SBP and DBP. FHS, Framingham Heart Study; SBP, systolic blood pressure; DBP, diastolic blood pressure.
